# Explaining COVID-19 Outbreaks with Reactive SEIRD Models

**DOI:** 10.1101/2021.02.09.21251440

**Authors:** Kunal Menda, Lucas Laird, Mykel J. Kochenderfer, Rajmonda S. Caceres

## Abstract

COVID-19 epidemics have varied dramatically in nature across the United States, where some counties have clear peaks in infections, and others have had a multitude of unpredictable and non-distinct peaks. In this work, we seek to explain the diversity in epidemic progressions by considering an extension to the compartmental SEIRD model. The model we propose uses a neural network to predict the infection rate as a function of time and of the prevalence of the disease. We provide a methodology for fitting this model to available county-level data describing aggregate cases and deaths. Our method uses Expectation-Maximization in order to overcome the challenge of partial observability—that the system’s state is only partially reflected in available data. We fit a single model to data from multiple counties in the United States exhibiting different behavior. By simulating the model, we show that it is capable of exhibiting both single peak and multi-peak behavior, reproducing behavior observed in counties both in and out of the training set. We also numerically compare the error of simulations from our model with a standard SEIRD model, showing that the proposed extensions are necessary to be able to explain the spread of COVID-19.

## 1. Introduction

Having an accurate understanding of the spread of COVID-19 is essential to be able to effectively contain the virus, and necessary for the deployment and allocation of resources. In order to understand why the spread appears to differ between communities, we require a mathematical model of disease spread capable of expressing the differences. A canonical model of disease spread is the SEIRD model, in which each individual is either susceptible to (S), exposed to (E), infected by (I), recovered from (R), or who have died from (D) the disease (Hethcote, 2000; Kermack and McKendrick, 1927). *Compartmental* SEIRD models consider only the aggregate number of individuals with each disease state, and specify a set of differential equations that govern how the compartmental populations change with time.

The well-studied compartmental SEIRD model is popular because of its simplicity (Hethcote, 2000). The standard model, however, predicts a single, clear peak in infections. While certain counties in the northeast of United States appeared to exhibit this behavior for a long duration, most other counties do not.

In this work, we seek to fit disease models to data that can account for the diversity in COVID-19 outbreaks across the US. Specifically, we seek to learn a model that predicts clear peaks for counties in the northeast that had them, and predicts multiple, flatter peaks for those that did not.

In order to build models capable of expressing more realistic behavior, we relax two assumptions made by the standard compartmental SEIRD model:

(A1) Stationarity—the disease parameters remain constant over time, and,

(A2) Non-reactivity—the disease parameters remain constant regardless of the prevalence of the disease.

We hypothesize that relaxing these assumptions allows us to better explain the diversity of behavior seen in reality.

To relax these assumptions, we consider a “reactive” compartmental SEIRD model (R-SEIRD) in which the transmission rate is a function of both time and of the number of infected individuals. To avoid imposing an incorrect prior on the functional form of this relationship, we use a neural network to model it.

A common difficulty in fitting such models to available data is that of *partial observability*. SEIRD models have *states* that vary with time according to their dynamic parameters. Because the available data are not time-series of the models’ states, but only partial observations of it (daily new cases and deaths), we employ tools from system-identification (Ljung, 1999) in order to fit these models. Specifically, we use a technique called Certainty-Equivalent Expectation Maximization (CE-EM) (Menda et al., 2020), which was recently shown to be a reliable and low-variance method for learning the parameters of partially observable dynamical systems.

In this work, we present a methodology for fitting R-SEIRD models to available data, and validate our hypotheses by showing that learned models can explain the diversity of behavior seen across the United States. Through our experiments, we show that:

1. Learned R-SEIRD models with appropriate initial conditions, and when simulated determistically, produce behavior consistent with what was observed in counties across the United States, and,
2. The simulation error of R-SEIRD models is lower than that of standard SEIRD models when compared against trajectories from outbreaks across the United States.

The contributions of this work are therefore to (1) propose a model that relaxes the assumptions of the standard SEIRD model, (2) provide a methodology for fitting this model to available data, and (3) experimentally demonstrate that this model is capable of expressing the diversity of behavior observed across the United States.

This paper is organized as follows. In Section 2, we formalize compartmental SEIRD models and review the CE-EM algorithm, as well as related approaches to fitting such models to data. In Section 3, we introduce the R-SEIRD model and describe our methodology for fitting it to available data. In Section 4, we fit the model to data from a selection of representative counties across the United States and show that it is able to reflect the observed behavior. Finally, in Section 5, we discuss possible limitations to the scope of our work, as well as directions for further study.

## 2 Background

In this section, we review compartmental SEIRD models, as well as CE-EM, an algorithm for fitting partially observed state-space models to data. We also discuss the literature addressing related problems.

### 2.1 SEIRD Models

SEIRD models are mathematical models of the spread of an infectious disease. Every individual in a population is in one of five states—they are either susceptible (S) to the disease, exposed (E) to the disease, infected (I) by the disease, or who have recovered (R) or died (D) from the disease (Hethcote, 2000). In this context, we assume an exposed individual is ‘pre-symptomatic’, i.e. they are able to spread the disease but have not yet tested positive for it, while infected individuals are symptomatic and have tested positive for the disease. There are numerous extensions to this model. For example, a SEIR model typically groups those who have recovered and died from the disease into a single state. SEIRD models may also differ in their modeling of reinfection, limited testing, or asymptomatic and quarantined individuals.

#### 2.1.1 Compartmental Models

In reality, each individual interacts with only a subset of individuals in the population, and thus the spread of the disease ought to be considered as propagating over a graph of sparsely connected nodes. However, by introducing this fidelity into the model, the state of the system exponentially grows with the number of individuals, and the task of fitting the model to data becomes difficult. A compartmental SEIRD model ignores the network structure of a population by making an assumption that the population is *homogeneously mixed*. That is, we assume that every individual in the population is equally likely to interact with any other individual in the population on a given day. By making this assumption, we can dramatically simplify the state of the system. We need only track the number of individuals in each disease state, referred to as the *compartmental populations*, and not specific individuals.

We can model the dynamics of the state *x*_*t*_ = [*S*_*t*_, *E*_*t*_, *I*_*t*_, (*RD*)_*t*_]^T^ as the following deterministic system of differential equations:

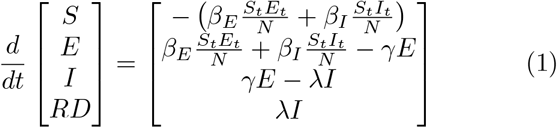

Here, the parameters *β*_*E*_ and *β*_*I*_ (referred to as the *infection rates*) are interpreted as the average number of individuals that an exposed or infected individual comes in contact with per unit time, multiplied by the probability that the contact results in disease transmission, respectively. The parameter *γ* specifies the average number of exposed individuals who transition to the infected state, per unit time, and *λ* specifies the average number of infected individuals who recover or die, per unit time. We define infected individuals to be those who have *tested positive* for the disease, and thus *γ*^*−*1^ is the average amount of time it takes for someone who contracted the disease to test positive for it. The *recovered-deceased* (RD) compartment includes both individuals who have truly recovered from the disease and those who have died from it. Hence, *λ*^*−*1^ is the average amount of time it takes for someone to no longer be infectious after having tested positive, as a result of recovery or death. If we assume a fixed mortality rate *µ* for the disease, then we can compute the number of individuals who have died from the disease as *D*_*t*_ = *µ*(*RD*)_*t*_, and those who have recovered from it as *R*_*t*_ = (1 − *µ*)(*RD*)_*t*_.

The above framing describes a deterministic dynamical system. There are many methods for framing compartmental SEIR systems using stochastic differential equations (Greenwood and Gordillo, 2009), and one such formulation will be discussed in Section 3.

It is worth noting the various effects *not* modeled here. In this model, we do not account for individuals who never develop symptoms. Furthermore, this model assumes an individual’s infectiousness changes when they test positive—the truth of which may depend on the delays experienced with RT-PCR testing. SEIR models can be extended to account for these effects, though such extensions may introduce more parameters and not be identifiable from the available data.

It would be straightforward to learn the parameters *θ* = [*β*_*E*_, *β*_*I*_, *γ, λ, µ*] from data if we could directly observe the compartmental populations. However, the available data is typically only of the aggregate transitions from the *E* compartment to the *I* compartment, when individuals test positive for the disease, and aggregate transitions from *I* to *R*, when individuals recover from or die due to the disease. As a result, estimation of *θ* must be performed under partial observability. Methods for doing so, including CE-EM, will be discussed next.

### 2.2 Certainty-Equivalent Expectation Maximization

Suppose we seek to find the parameters of the following state-space dynamical system:

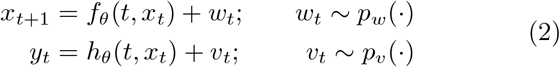

Here, *x*_*t*_ is the state of the system, *f*_*θ*_(*t, x*_*t*_) is a parameterized model of the dynamics, *w*_*t*_ is an additive *process noise* term, and *p*_*w*_ is the distribution from which *w*_*t*_ is sampled. Furthermore, *y*_*t*_ is an observation of *x*_*t*_, *h*_*θ*_(*t, x*_*t*_) is a parameterized observation model, *v*_*t*_ is the *observation noise* term, and *p*_*v*_ is the distribution from which *v*_*t*_ is sampled. In a *nonlinear Gaussian* system, *p*_*w*_ and *p*_*v*_ are multivariate Gaussian distributions.

Methods for parameter estimation typically attempt to quantify the likelihood *p*(*y*_1:*T*_ | *θ*) of a time-series of observations *y*_1:*T*_ given some choice of parameters *θ*. A set of methods called approximate Bayesian computing (ABC) use this estimate to characterize the Bayesian posterior *p*(*θ | y*_1:*T*_) using approximate methods such as Markov Chain Monte Carlo (Brown et al., 2018; Sunn°aker et al., 2013). Maximum-likelihood (MLE) methods attempt to find the likelihood maximizing parameters *θ*_ML_ = arg max *p*(*y*_1:*T*_ |*θ*). Both methods rely on being able to estimate the data likelihood *p*(*y*_1:*T*_ | *θ*) with low-variance.

In order to estimate the data likelihood, we must marginalize over the unobserved states of dynamical system.

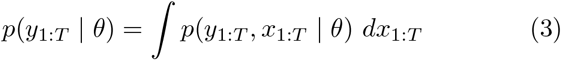

Many approaches to fitting SEIR models to data assume known initial conditions *x*_1_ when estimating *p*(*y*_1:*T*_ *θ*) (Gu, 2020; He et al., 2020; Korolev, 2021). Key drawbacks to such approaches are their sensitivity to the choice of *x*_1_, as well as a degradation in their ability to find global optima if being fit to long timeseries. Since computing this expectation is generally intractable without knowing initial conditions and for long time-series, many approaches instead find the posterior distribution over states, i.e. *p*(*x*_1:*T*_ | *y*_1:*T*_, *θ*), and then approximate quantities of interest using Monte-Carlo samples (Kantas et al., 2015).

In a subset of MLE methods, the *smoothing distribution* distribution *p*(*x*_1:*T*_ | *y*_1:*T*_, *θ*) is used to estimate the joint data log-likelihood:

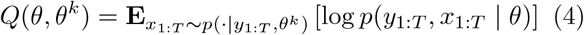

The function *Q*(*θ, θ*^*k*^) is then maximized to yield *θ*^*k*+1^, and this two-step procedure is repeated until convergence, in an algorithm called Expectation-Maximization (EM) (Dempster et al., 1977). Such approaches overcome the need to precisely know *x*_1_ and can straightforwardly handle long time-series (Menda et al., 2020).

Various algorithms differ in how they compute the smoothing distribution. ParticleEM uses particle smoothing, an approach that uses sequential Monte-Carlo to approximately sample from the smoothing distribution (Kantas et al., 2015; Schön et al., 2011). Though this approach makes very few assumptions, it can require prohibitive number of Monte-Carlo samples to yield sufficiently low variance estimates of *Q*(*θ, θ*^*k*^).

The Certainty-Equivalent EM (CE-EM) algorithm makes the approximation that *p*(*x*_1:*T*_ | *y*_1:*T*_, *θ*) can be modeled by a Dirac-delta function located at the smoothing distribution’s mode (Menda et al., 2020). In the case where *p*_*w*_(*·*) and *p*_*v*_(*·*) are Gaussian distributions, CE-EM can efficiently find an estimate for *θ*^ML^ with low-variance using block-coordinate ascent. However, by making this approximation, the algorithm is known to be biased in the presence of large process noise.

In this work, we propose to use CE-EM to fit SEIRD models, including our proposed extension to them, to available county-level data. As one might expect, such an approach should yield inaccurate results if the disease progression is highly stochastic, or poorly modeled by a compartmental model. However, we will show that even when fitting a single model to data from six counties with diverse epidemics, simulations from the model appear to reflect reality.

### 2.3 Related Work

Since the dawn of the COVID-19 pandemic, many efforts have been directed at forecasting its evolution, as well as fitting modified SEIR models to specific outbreaks (Ahmad et al., 2020; Naudé, 2020). Approaches vary in their characterization of the model, and their methodology for parameter estimation under partial observability. Korolev (2021) demonstrates issues with the identifiability of the SEIRD model and presents an estimation technique for the basic reproduction number *R*_0_. They fit a standard SEIRD model to COVID-19 data but assume known initial conditions to address partial observability—an assumption that can only made in restricted settings. He et al. (2020) use particle swarm optimization to optimize the parameters of a SEIR model extended to account for hospitalized and quarantined individuals. They also address partial observability by assuming known initial conditions, but only fit to a single, short trajectory of data from the Hubei province in China. Arik et al. (2020) introduce additional data sources such as mobility as covariates to expand the explanatory power of SEIR models, and specify a distribution over initial conditions to overcome partial observability. One of the more successful approaches to forecasting the early pandemic progression also fit SEIR models with a subset of variables allowed to vary with time (Gu, 2020). Their approach also assumes a known initial condition, and finds parameters by minimizing simulation error.

Recent works (Dandekar and Barbastathis, 2020; Melin et al., 2020; Wieczorek et al., 2020) have also attempted to use neural networks for the purpose of forecasting the spread of COVID-19. While Melin et al. (2020); Wieczorek et al. (2020) do not use neural-networks to model relationships within a SEIR model, Dandekar and Barbastathis (2020) use them to model a quarantine control function. Similar to other discussed methodologies, they assume initial conditions to overcome partial observability. Yang et al. (2020) fit both a SEIR model with additional compartments, as well as a black-box LSTM to a short time-series of COVID-19 infection data from Hubei, China, and compare their forecasting ability. Their SEIR model is not fit to the data by attempting to reproduce a time-series, but by linearizing the model around certain set points and assuming values for unknown quantities.

To our knowledge, our work is the first to use a neural network to model the relationship between time, prevalence, and infection rate, the first to use Certainty-Equivalent EM for the purpose of parameter estimation under partial observability, and also the first to fit a single model to multiple time-series from across the United States.

## 3 Methodology

In this section, we describe data sources used in this work, the proposed modification to SEIRD models yielding R-SEIRD models, the formulation of R-SEIRD models as nonlinear Gaussian systems, and a procedure for fitting these models to available data.

### 3.1 Data Sources

In this work, the primary data source considered is the Novel Coronavirus (COVID-19) Cases Dataset, provided by JHU CSSE (Dong et al., 2020), which provides a time-series of daily reported COVID-19 cases and deaths for every county in the United States.

### 3.2 R-SEIRD Models

As stated in Section 1, we propose to relax that the assumption that the infection rate *β*_*E*_ is constant with time. Instead, we model it to be a function of time and the *observed prevalence* of the disease. Specifically, we let:

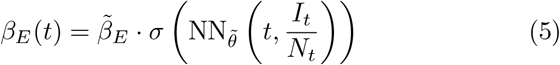

Here, *N*_*t*_ = *S*_*t*_ + *E*_*t*_ + *I*_*t*_ + (*RD*)_*t*_ is the *effective* population size at time *t*, a quantity that we allow to vary with time (Li et al., 1999) and be dynamically inferred during learning. Additionally, *I*_*t*_*/N*_*t*_ is the observed prevalence, 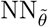 is a neural network mapping **R**^2^ *→* **R**, *σ*(*·*) is the sigmoid function, and 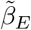 is a learned coefficient. For simplicity, we assume *β*_*I*_ = 0, which implies that the number of infections caused by an individual after they test positive for COVID-19 is negligible compared to the number of infections prior to them knowing they have the disease. Furthermore, since much is now known about the typical duration between exposure and symptom onset, and symptom onset and death, we rely on literature to provide estimates of *γ* and *λ* (Bi et al., 2020; Lauer et al., 2020). Our methodology does, however, allow us to treat them as learned parameters. We treat the mortality rate *µ* as learned, and thus 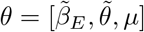 are the learned parameters.

With this modification, we allow the model to reflect changes in the infection rate that may depend on changing behaviors over time, season, and in response to the level of infection in a county.

### 3.3 R-SEIRD as a Nonlinear Gaussian System

In order to effectively learn the parameters of an R-SEIRD model using CE-EM, we represent it as a non-linear Gaussian system. To do so, we must specify the state *x*_*t*_, discrete-time dynamics function *f*_*θ*_(*t, x*_*t*_), observation *y*_*t*_, and observation function *h*_*θ*_(*t, x*_*t*_).

Though the state of a SEIRD system is described by the number of individuals in each compartment, we note two facts about these quantities:

- The number of individuals in any compartment is a positive quantity, and,
- These quantities scale various orders of magnitude (Dong et al., 2020).

For this reason, it is sensible to let the state *x*_*t*_ of the system correspond to the logarithm of each compartment’s population as opposed to their absolute value. As a result, applying Gaussian process or observation noise to these quantities loosely corresponds to assuming that noise is proportional to the absolute value of the quantity it is applied to. Specifically, let the state of the system be *x*_*t*_ = log [*S*_*t*_, *E*_*t*_, *I*_*t*_, (*RD*)_*t*_], and the observation be *y*_*t*_ = log [Δ*C*_*t*_, Δ*D*_*t*_]. Here, Δ*C*_*t*_ corresponds to the number of new confirmed cases on day *t*, and tracks the total number of individuals that have transitioned from the *E* to the *I* compartment, and Δ*D*_*t*_ corresponds to the number of new deaths on day *t*.

The dynamics and observation models for the R-SEIRD model are then specified as follows:

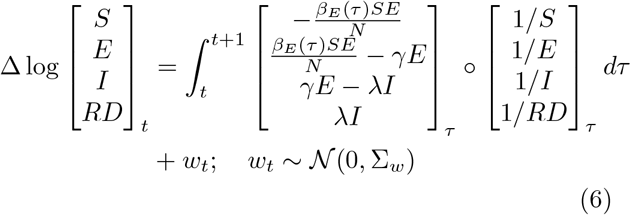

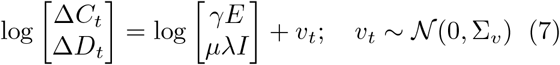

where *β*_*E*_(*t*) is computed according to Equation (5). The process noise covariance Σ_*w*_ and observation noise covariance Σ_*v*_ are hyperparameters that can be chosen to be identity or diagonal matrices scaled by 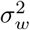 and 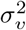 respectively. Integration is performed using a Runge-Kutta method (Runge, 1895).

Since the parameters 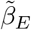 and *µ* are positive, we optimize their logarithms as opposed to their absolute values. Framed as a nonlinear Gaussian system, the R-SEIRD model can be straightforwardly fit to a batch of observation time-series from multiple counties simultaneously by using CE-EM (Menda et al., 2020). To improve fit reliability and eliminate periodic drops from weekends, we apply a 7-day moving average filter to daily case and death data before fitting to them.

In our experiments, we fit the R-SEIRD model to data from six counties in the United States exhibiting diversity in how COVID-19 has spread, and show that it is capable of expressing this diversity.

## 4 Experiments

Our experiments aim to achieve the following goals:

1. Fit a single R-SEIRD model to data from a variety of counties across the United States,
2. Show that the learned model, when simulated with realistic initial conditions, is able to reproduce observed behavior, and,
3. Justify the modifications to the SEIRD model by showing that the mean squared error (MSE) of simulations under R-SEIRD model is much lower than that of a fit SEIRD model.

To achieve these goals, we fit models to data from three counties in the northeast of the United States (Middlesex, MA, Kings, NY, and Fairfield, CT), as well as three counties across the United States that have had diverse epidemics (Los Angeles, CA, Miami-Dade, FL, and Cook, IL). We consider data from the February 22, 2020 to the September 27, 2020, because during this period the counties in the northeast exhibit single, clear peaks in infection, though the remaining counties exhibit multiple peaks. The remaining worst-hit counties in each of the United States compose a test-set to evaluate the model.

We fit both an R-SEIRD model and a standard SEIRD model (where we learn just the constant parameters *β*_*E*_ and *µ*) to data from these six counties, and then attempt to see if the learned models can reproduce the behavior of all six counties, as well as of counties not trained on, from appropriate initial conditions. We visually compare simulations on select counties to show that deterministic simulations of the R-SEIRD system are capable of expressing multiple peaks while those of a SEIRD system are not. We also visualize the learned relationship between time, prevalence, and infection rate, to provide an intuition for how the model is able to express such behavior. We then compare MSE of simulations from both learned models on the worst-hit county of each of the United States (not in the training set), showing that simulation error is much lower when using the R-SEIRD model^[1]^.

### 4.1 Experimental Setup

Here we detail the hyperparameters used when training the R-SEIRD and SEIRD models, the methodology for selecting appropriate initial conditions for states when evaluating the models, as well as metrics for evaluation.

#### 4.1.1 Hyperparameters

When learning the R-SEIRD model or SEIRD model,

we specify the values of *γ* and *λ*. The literature suggests that the median time from exposure to developing symptoms is five days (Lauer et al., 2020) and that the median time between symptom onset and recovery is 21 days (Bi et al., 2020). As a result, we let *γ* = 1*/*5 and *λ* = 1*/*21. We let *σ*_*w*_ = *σ*_*v*_*/*10, assuming that corruptions to observations are an order of magnitude noisier than corruptions to the process. Furthermore, we let the observation noise on deaths be double that on cases, since reporting of deaths is typically more noisy when the number of deaths is small. The neural network used in the R-SEIRD model has three hidden layers with 32 hidden units each and TanH activation functions. Both models are optimized in the CE-EM learning step using an Adam optimizer with a learning rate of 5 *×* 10^*−*4^, which is harmonically decayed over time, and CE-EM trust-region parameters of *ρ*_*x*_ = 0.5 and *ρ*_*θ*_ = 0.01 (Menda et al., 2020).^[2]^

#### 4.1.2 Initial Condition Selection

When evaluating learned models, partial observability makes it such that we do not know the initial conditions (i.e. *x*_1_) to simulate the models from. Related work often assumes the population is almost completely unexposed at *t* = 1, and assumes the susceptible population is a community’s true population (He et al., 2020). However, as stated in Section 3, we allow the population size to be determined by the sum of the compartmental population, and therefore be a free parameter, and also work with trajectories in which it is unreasonable to assume an unexposed initial population.

For these reasons, we optimize for the initial conditions from which deterministically simulated trajectories minimize the MSE of the observed number of daily cases. We optimize the initial conditions by using the Cross Entropy Method (Rubinstein, 1999), selecting the top 100 candidates at each epoch, perturbing each candidate to generate 1000 candidates for the next epoch, and running the optimizer for 10 epochs.

#### 4.1.3 Evaluation Metrics

When selecting initial conditions, we measure the MSE between the log of the daily cases in simulation (i.e. log(*γE*_*t*_)), and reality (i.e. log Δ*C*_*t*_), and use the distribution over this metric to compare the R-SEIRD and standard SEIRD models. We compare histograms of these errors on data from counties not in the training set.

#### 4.1.4 Results

In Figure 1, we see the simulated number of cases from optimized initial conditions for the six counties on which the R-SEIRD model was trained. As is clearly seen, the model is capable of expressing both a single clear peak present in the northeastern counties, as well as the multiple irregular peaks present in the other counties. Since the simulations are deterministic, they are smooth and only approximately reflect the trends in the trajectories they are supposed to match, but do not reflect the volatility in the real data.

**Figure 1:**
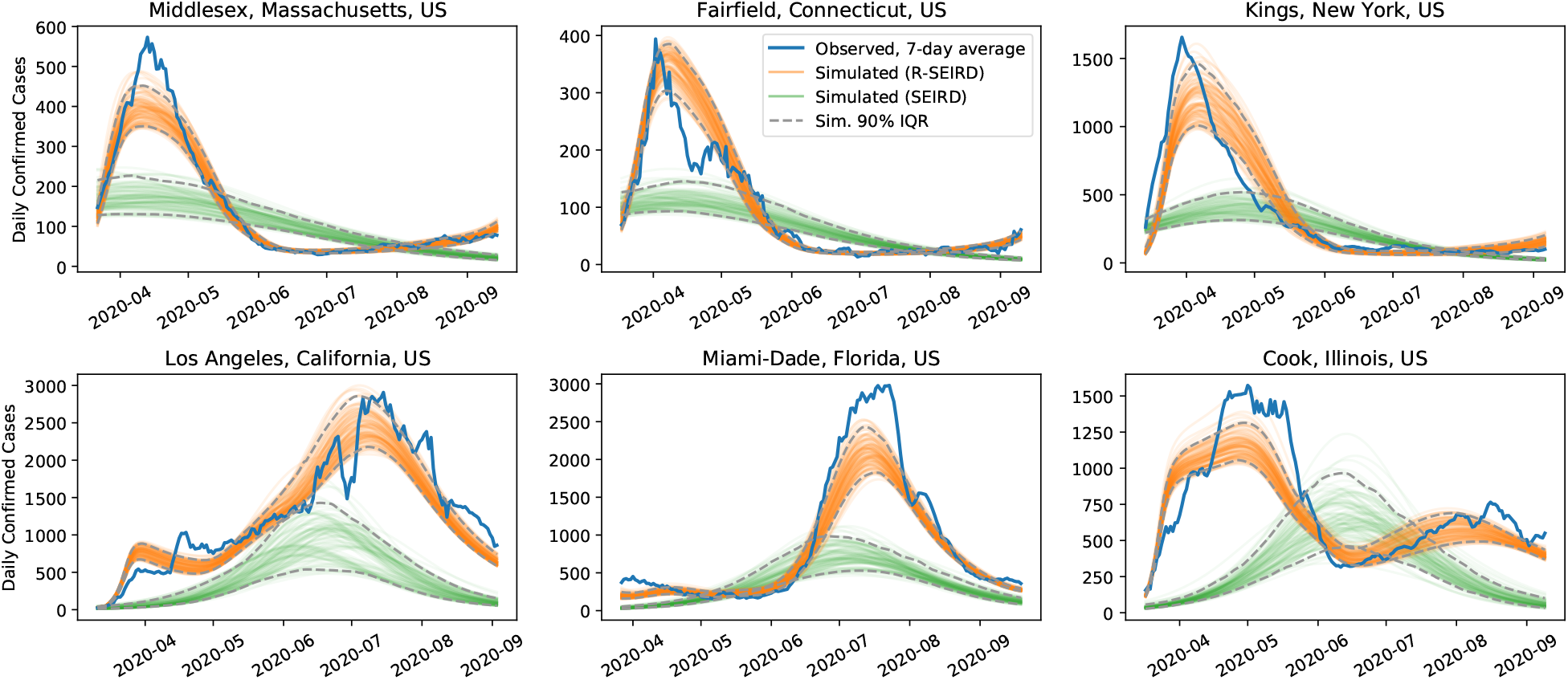
Simulations of the learned R-SEIRD and learned SEIRD systems on the six counties the models are trained on.

In contrast, and consistent with expectation, a SEIRD model fit to the same data and simulated from optimized initial conditions is incapable of reflecting the multiple peaks in the training data. Furthermore, in an attempt to fit the multi-peak behavior in three of the counties, the fit is severely compromised in counties in which there is a single, clear peak.

In Figure 2, we compare the learned R-SEIRD and SEIRD models on three hard-hit counties not in the training set. Again, the SEIRD model is unable to express more than a single peak, and thus is not able to capture the behavior reflected in reality. Despite not being trained on data from these counties, the R-SEIRD model is, however, able to reflect the double-peaks seen in the data, which vary in onset and magnitude.

**Figure 2:**
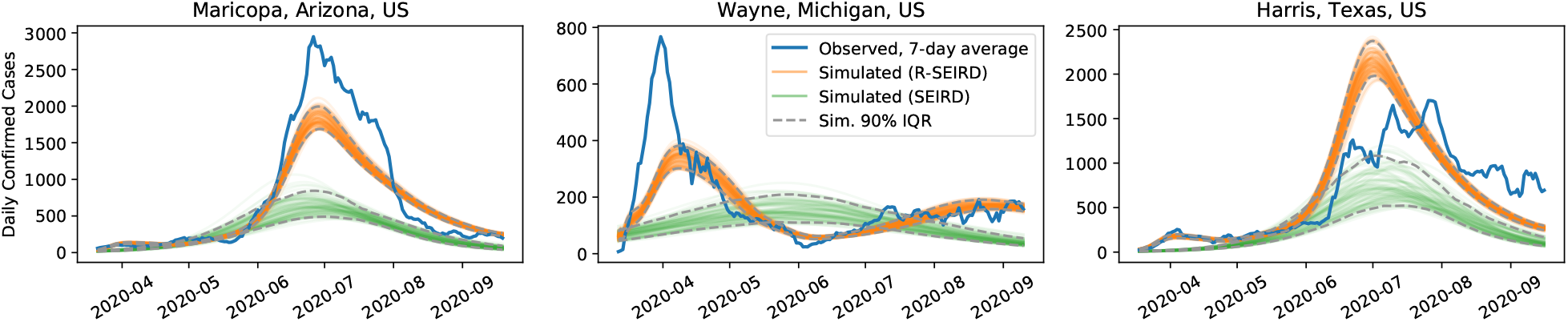
Simulations of the learned R-SEIRD and learned SEIRD systems on the three counties the models are not trained on.

We quantify the difference in simulation quality by measuring the MSE between the simulations (from optimized initial conditions) with the observed trajectories. In Figure 3, we show a histogram of these errors for simulations on the worst-hit county of each of the United States (not in the training set). Consistent with the improvement visible in Figure 2, we see that the MSE of simulations by the learned R-SEIRD model is significantly lower than that of the SEIRD model.

**Figure 3:**
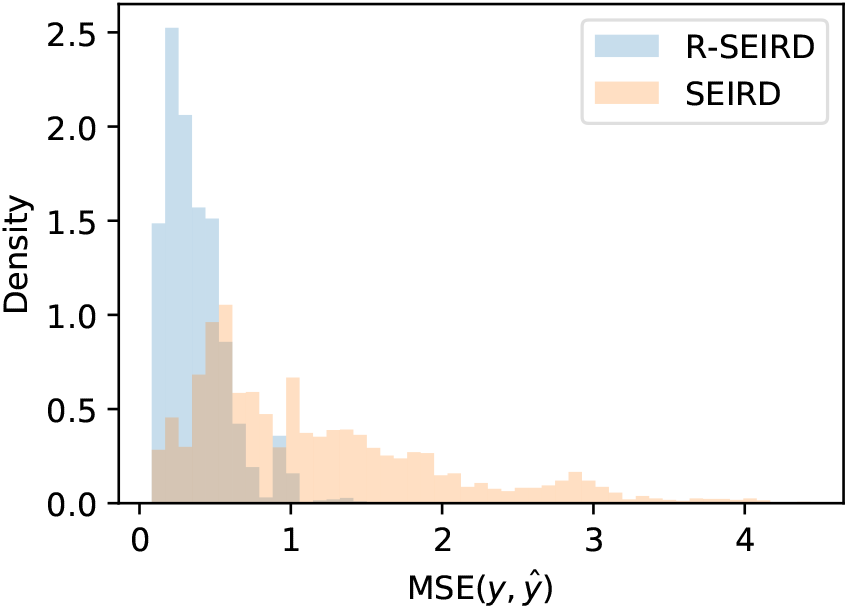
Comparison of prediction errors between R-SEIRD and SEIRD for the epidemic in the worst-hit county of each of the United States.

To gain an intuition for how the R-SEIRD model is able to express multiple irregular peaks, we visualize the learned mapping between time, the prevalence of the disease (*I*_*t*_*/N*_*t*_) and the basic reproduction number *R*_0_ = *β*_*E*_(*t, I*_*t*_*/N*_*t*_)*/γ*, which indicates an growing epidemic for values greater than 1. In Figure 4, we see the learned mapping, which suggests that *β*_*E*_ deceases initially as prevalence increases, up until a point, after which it begins to surge again before decreasing. This behavior would be a consistent with a two-tiered response, in which weak restrictions are adopted until they lose effect, and then stronger restrictions are imposed. Over time, we see that *β*_*E*_ begins to require larger levels of prevalence to decay and bring *R*_0_ below one, which could suggest that communities are growing more reluctant to adopt the weak restrictions when infection rates are low.

**Figure 4:**
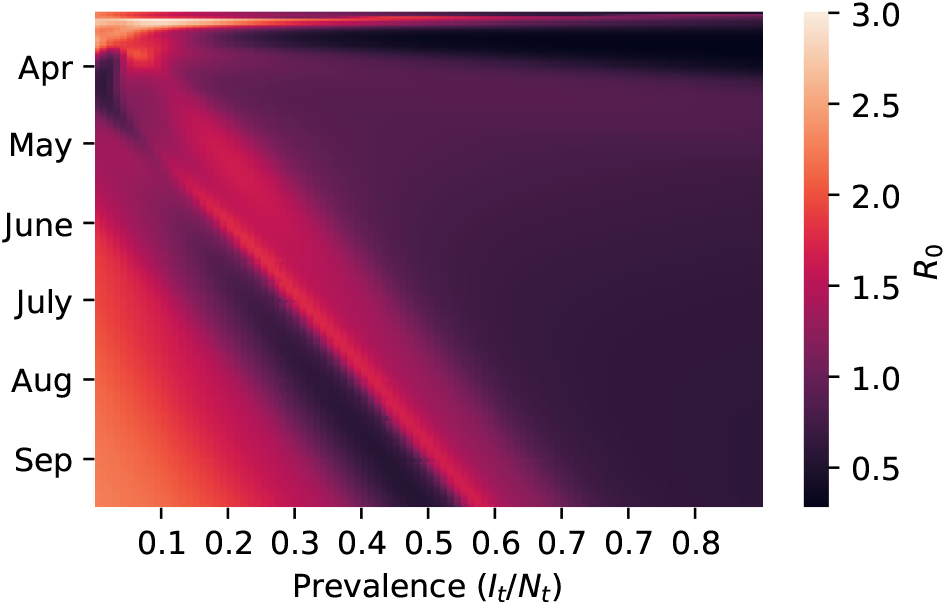
The learned mapping between time and prevalence to the infection rate.

## 5 Conclusion

In this work, we proposed an extension to the compartmental SEIRD model that relaxes the assumptions of stationarity and non-reactivity called the R-SEIRD model. We did so by training a neural network to map the time and the prevalence of the disease to the infection rate. In order to fit to available data, we employed Certainty-Equivalent Expectation Maximization (CE-EM), which a technique suited to fitting nonlinear Gaussian state-space models to data without direct observation of the system’s state variables. We provided a methodology for framing the R-SEIRD model as a nonlinear Gaussian system, and for fitting it to available data on daily confirmed cases and deaths using CE-EM.

In our experiments, we fit both the R-SEIRD and standard compartmental SEIRD models to data from six counties across the United States. We showed that the R-SEIRD model learned is capable of expressing the range of multi-peak behavior exhibited not only in the training data, but also on counties not trained on. We showed quantitatively that the simulation error when trying to reproduce the behavior of the worst-hit counties in the United States is much lower when using the R-SEIRD model, when compared to the standard SEIRD model.

This work showed that by allowing the infection rate to be time-varying and reactive, the range of behaviors exhibited by the epidemic across the United States can be recovered. We do not, however, suggest that the R-SEIRD model definitively explains why a given outbreak progressed the way it did, but rather proposes a hypothesis that is consistent with observations. To come closer to a definitive explanation, we must depart from another assumption made by both standard compartmental SEIRD and R-SEIRD models—that of *homogeneous mixing*. By modeling a counties’ populations as individuals connected by a topological *graph*, we gain the capacity to account super-spreading individuals, as well as non-uniform population densities.

In a follow-on to this work, we aim to propose a SEIRD model that accounts for the network structure of a community, as well as an estimation procedure for fitting it to data. With such a model, we not only gain another explanatory tool to analyze the diversity in outbreaks, but also a model that can be used to evaluate localized containment strategies at a community level.

## Data Availability

The datasets generated and/or analyzed during the current study are available in the COVID-19 Data Repository by the Center for Systems Science and Engineering (CSSE) at Johns Hopkins University (https://github.com/CSSEGISandData/COVID-19)

https://github.com/sisl/rseird

## Acknowledgments

The authors would like to acknowledge Neela Kaushik, Ross Alexander, Christian Vanderloo, and Shushman Choudhury for their valuable insights.

## Funding

This material is supported by the Under Secretary of Defense for Research and Engineering under Air Force Contract No. FA8702-15-D-0001. Any opinions, findings, conclusions or recommendations expressed in this material are those of the author(s) and do not necessarily reflect the views of the Under Secretary of Defense for Research and Engineering.

### Abbreviations

CE-EM: Certainty-Equivalent Expectation-Maximization. 2
R-SEIRD: Reactive-SEIRD. 1
SEIRD: Susceptible, Exposed, Infected, Recovered, Deceased. 1

## Availability of data and materials

The datasets generated and/or analyzed during the current study are available in the COVID-19 Data Repository by the Center for Systems Science and Engineering (CSSE) at Johns Hopkins University (https://github.com/CSSEGISandData/COVID-19) (Dong et al., 2020).

## Authors’ contributions

Kunal Menda proposed the methodology for this manuscript and conducted the experiments. All authors contributed to writing of the manuscript and the review of relevant literature.

## STROBE Statement—checklist of items that should be included in reports of observational studies

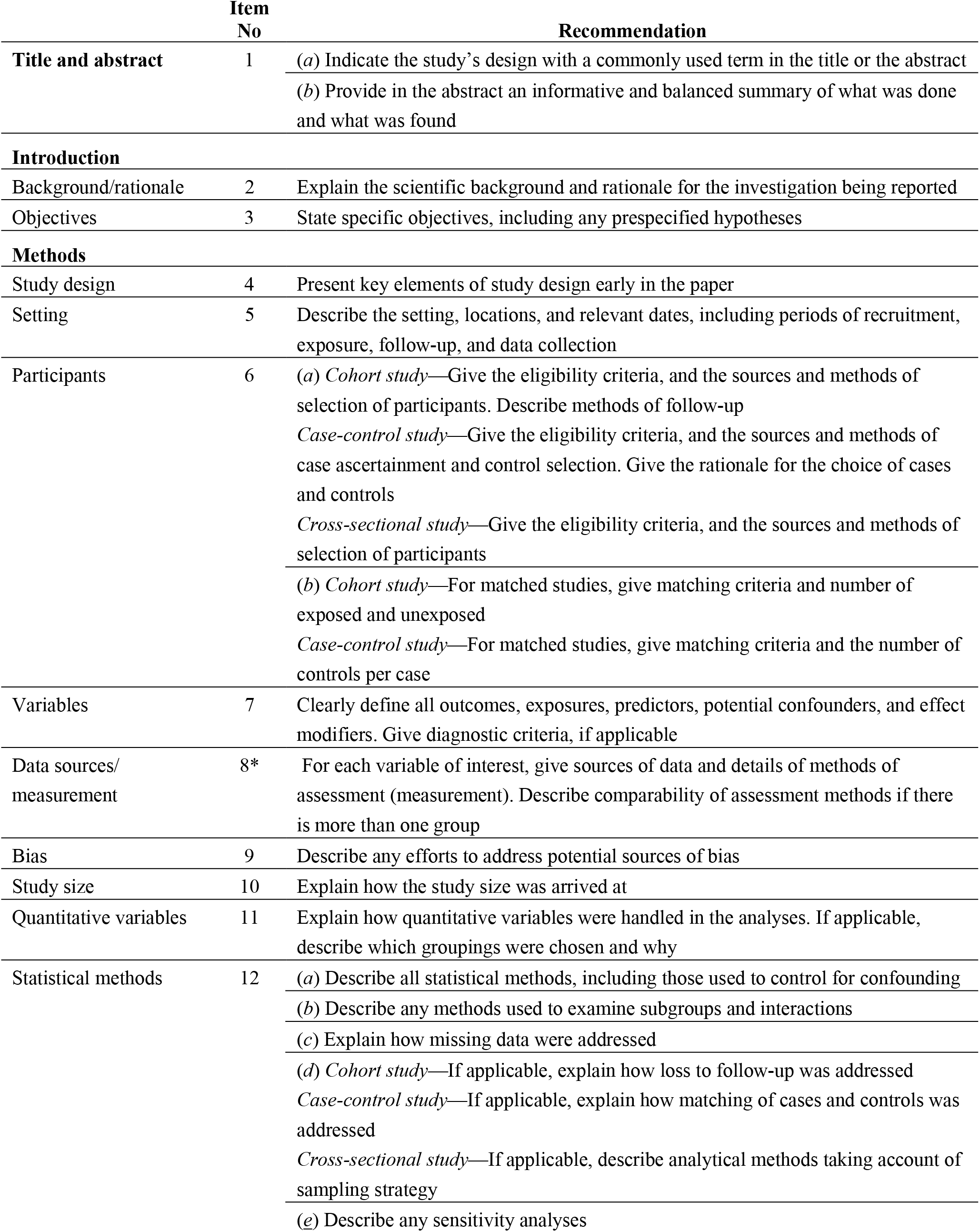
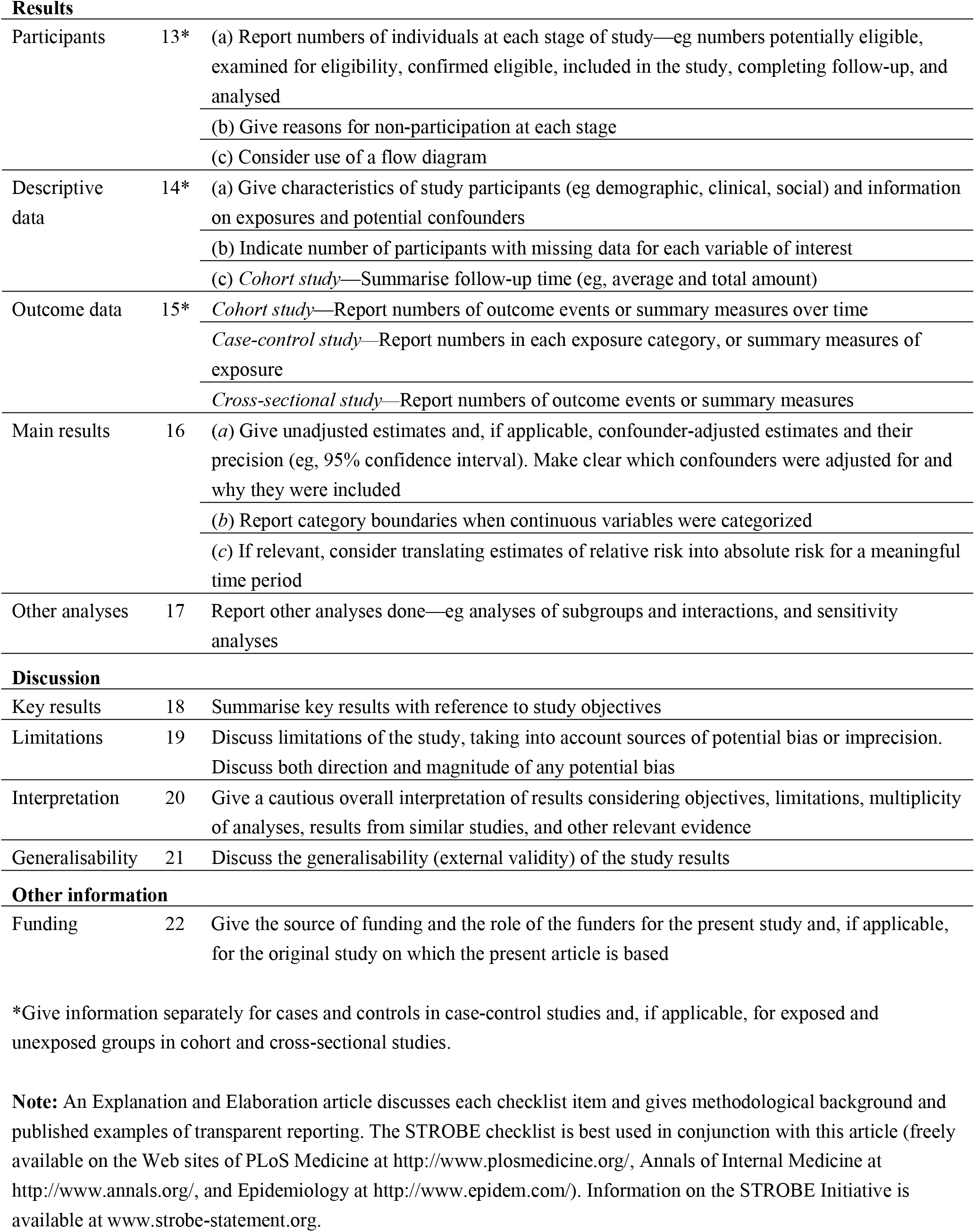

|  | Item No | Recommendation |
| --- | --- | --- |
| **Title and abstract** | 1 | (a) Indicate the study's design with a commonly used term in the title or the abstract<br>(b) Provide in the abstract an informative and balanced summary of what was done and what was found |
| **Introduction** |  |  |
| Background/rationale | 2 | Explain the scientific background and rationale for the investigation being reported |
| Objectives | 3 | State specific objectives, including any prespecified hypotheses |
| **Methods** |  |  |
| Study design | 4 | Present key elements of study design early in the paper |
| Setting | 5 | Describe the setting, locations, and relevant dates, including periods of recruitment, exposure, follow-up, and data collection |
| Participants | 6 | (a) *Cohort study*—Give the eligibility criteria, and the sources and methods of selection of participants. Describe methods of follow-up<br>*Case-control study*—Give the eligibility criteria, and the sources and methods of case ascertainment and control selection. Give the rationale for the choice of cases and controls<br>*Cross-sectional study*—Give the eligibility criteria, and the sources and methods of selection of participants<br>(b) *Cohort study*—For matched studies, give matching criteria and number of exposed and unexposed<br>*Case-control study*—For matched studies, give matching criteria and the number of controls per case |
| Variables | 7 | Clearly define all outcomes, exposures, predictors, potential confounders, and effect modifiers. Give diagnostic criteria, if applicable |
| Data sources/<br>measurement | 8* | For each variable of interest, give sources of data and details of methods of assessment (measurement). Describe comparability of assessment methods if there is more than one group |
| Bias | 9 | Describe any efforts to address potential sources of bias |
| Study size | 10 | Explain how the study size was arrived at |
| Quantitative variables | 11 | Explain how quantitative variables were handled in the analyses. If applicable, describe which groupings were chosen and why |
| Statistical methods | 12 | (a) Describe all statistical methods, including those used to control for confounding<br>(b) Describe any methods used to examine subgroups and interactions<br>(c) Explain how missing data were addressed<br>(d) *Cohort study*—If applicable, explain how loss to follow-up was addressed<br>*Case-control study*—If applicable, explain how matching of cases and controls was addressed<br>*Cross-sectional study*—If applicable, describe analytical methods taking account of sampling strategy<br>(e) Describe any sensitivity analyses |
| **Results** |  |  |
| Participants | 13* | (a) Report numbers of individuals at each stage of study—eg numbers potentially eligible, examined for eligibility, confirmed eligible, included in the study, completing follow-up, and analysed<br>(b) Give reasons for non-participation at each stage<br>(c) Consider use of a flow diagram |
| Descriptive data | 14* | (a) Give characteristics of study participants (eg demographic, clinical, social) and information on exposures and potential confounders<br>(b) Indicate number of participants with missing data for each variable of interest<br>(c) *Cohort study*—Summarise follow-up time (eg, average and total amount) |
| Outcome data | 15* | *Cohort study*—Report numbers of outcome events or summary measures over time<br>*Case-control study*—Report numbers in each exposure category, or summary measures of exposure<br>*Cross-sectional study*—Report numbers of outcome events or summary measures |
| Main results | 16 | (a) Give unadjusted estimates and, if applicable, confounder-adjusted estimates and their precision (eg, 95% confidence interval). Make clear which confounders were adjusted for and why they were included<br>(b) Report category boundaries when continuous variables were categorized<br>(c) If relevant, consider translating estimates of relative risk into absolute risk for a meaningful time period |
| Other analyses | 17 | Report other analyses done—eg analyses of subgroups and interactions, and sensitivity analyses |
| **Discussion** |  |  |
| Key results | 18 | Summarise key results with reference to study objectives |
| Limitations | 19 | Discuss limitations of the study, taking into account sources of potential bias or imprecision. Discuss both direction and magnitude of any potential bias |
| Interpretation | 20 | Give a cautious overall interpretation of results considering objectives, limitations, multiplicity of analyses, results from similar studies, and other relevant evidence |
| Generalisability | 21 | Discuss the generalisability (external validity) of the study results |
| **Other information** |  |  |
| Funding | 22 | Give the source of funding and the role of the funders for the present study and, if applicable, for the original study on which the present article is based |

\*Give information separately for cases and controls in case-control studies and, if applicable, for exposed and unexposed groups in cohort and cross-sectional studies.
**Note:** An Explanation and Elaboration article discusses each checklist item and gives methodological background and published examples of transparent reporting. The STROBE checklist is best used in conjunction with this article (freely available on the Web sites of PLoS Medicine at http://www.plosmedicine.org/, Annals of Internal Medicine at http://www.annals.org/, and Epidemiology at http://www.epidem.com/). Information on the STROBE Initiative is available at www.strobe-statement.org.

## Footnotes

[1] The codebase for running these experiments can be found at https://github.com/sisl/rseird.

[2] [2] Experiments were run on 2.9 GHz Quad-Core Intel Core i7 MacBook Pro with 16 GB of RAM, and each model took under 15 minutes to train.

